# Development and evaluation of a kidney health questionnaire and estimates of chronic kidney disease prevalence in the Cooperative Health Research In South Tyrol (CHRIS) study

**DOI:** 10.1101/2024.03.24.24304607

**Authors:** Barbieri Giulia, Cazzoletti Lucia, Melotti Roberto, Hantikainen Essi, Lundin Rebecca, Barin Laura, Gögele Martin, Riegler Peter, Ferraro Pietro Manuel, Pramstaller Peter Paul, Gambaro Giovanni, Zanolin Maria Elisabetta, Pattaro Cristian

**Affiliations:** Institute for Biomedicine, Eurac Research, Via Volta 21, 39100, Bolzano/Bozen, Italy; Unit of Epidemiology and Medical Statistics, Department of Diagnostics and Public Health, University of Verona, Italy; Independent scholar, Bolzano/Bozen, Italy; Department of Medicine, Section of Nephrology, Department of Medicine, Università degli Studi di Verona, Verona, Italy

## Abstract

**Introduction:** Kidney diseases are a public health burden but, except for chronic kidney disease (CKD), they are poorly investigated in the general population. In light of inadequate survey tools for use in population studies, we developed a novel questionnaire to retrospectively assess the main kidney diseases, integrating it within the Cooperative Health Research In South Tyrol (CHRIS) study conducted between 2011 and 2018 in the Alpine district of Val Venosta/Vinschgau (Italy).

**Methods:** The questionnaire covers general kidney diseases, reduced kidney function, and renal surgeries. It was applied to the cross-sectional assessment of self-reported kidney health among 11,684 adults, along with measures of fasting estimated glomerular filtration rate (eGFR) and urinary albumin-to-creatinine ratio (UACR). By factor analysis we contextualized the questionnaire content with respect to biochemical measurements. CKD was defined according to KDIGO guidelines, self-reported diagnosis, and their combination. Prevalence estimates were calibrated to the general target population via relative sampling weights.

**Results:** In this population sample (median age=45 years; median eGFR=98.4 mL/min/1.73 m^2^; median UACR=5.7 mg/g), 8.3% of participants reported at least one kidney disease. Population-representative prevalence of glomerulonephritis, pyelonephritis, and congenital kidney diseases was 1.0%, 3.0%, and 0.2%, respectively, with corresponding odds ratio for females versus males of 1.4 (95% confidence interval: 1.0, 2.0), 8.7 (6.2, 12.3), and 0.7 (0.3, 1.6), respectively. CKD prevalence was 8.6% when based on KDIGO criteria and 0.7% when self-reported, indicating that 95.3% of affected individuals were unaware of having CKD, with a similar figure of 92.9% in those reporting diabetes or hypertension. Overall, 15.8% of the population was affected by a kidney disease of any kind.

**Conclusion:** In this Alpine population, CKD prevalence aligned with Western European estimates. Kidney health questionnaire implementation in population studies is feasible and valuable to assess CKD awareness, which we found to be dramatically low.

## Introduction

Chronic kidney disease (CKD) is becoming one of the leading causes of death worldwide.^1^ It affects individual health and quality of life, significantly burdening healthcare systems. With a global prevalence above 10%,^2^ and significant geographic variations both between-^3^ and within-continents,^4^ CKD’s distribution is influenced by environmental, behavioral, and genetic determinants, and public health policies.^4,5^ Hence, locally monitoring CKD prevalence is crucial in controlling this chronic disease.

In Italy, the latest estimates of CKD prevalence date back a decade. Based on estimated glomerular filtration rate (eGFR) and albuminuria measurements, CKD prevalence was estimated at 12.7% in 40+ year-old individuals^6^ and 7.1% in those 35-79 years old.^7^These figures indicated an increase compared to the previous decade, with age-adjusted CKD prevalence at 5.7% for males and 6.2% for females.^8^ The same study observed that less than 6% of the prevalent cases reported a diagnosis of any kidney disease. This gap between measured and self-reported CKD diagnosis aligns with the much lower standardized CKD prevalence (1.8%) estimated in the Lazio region (Italy) using a classification algorithm based on administrative data,^9^ reflecting the critical issue of CKD underdiagnosis.^10,11^

Besides CKD, other kidney diseases are relatively common, but lack robust prevalence estimates. These include glomerulonephritis, a primary factor in kidney failure,^12,13^ and pyelonephritis, a leading cause of hospitalization.^14^ Except for measured blood and urinary markers, tools for surveying CKD and other kidney-related conditions in the general population remain limited. For this reason, we designed a questionnaire to identify broad categories of kidney diseases for application to general population studies. The questionnaire was implemented in a sizeable central-European population study conducted in the Val Venosta/Vinschgau district (Italy), the Cooperative Health Research in South Tyrol (CHRIS) study.

Here, we present a comprehensive analysis of the questionnaire and its relationship with eGFR and albuminuria measured at the time of the interview. We provide population-standardized prevalence estimates for each type of kidney disease and different definitions of CKD, thus quantifying CKD awareness and underdiagnosis in the region.

## Methods

### Study design

At its baseline visit, the CHRIS study enrolled 13,388 consenting adults from the Val Venosta/Vinschgau district (South Tyrol, Italy) between 2011 and 2018.^15^ Following overnight fasting, participants underwent blood drawing, urine collection, blood pressure and anthropometric measurements, and clinical examinations. Participants filled out standardized questionnaires investigating demographics, medical history, and lifestyle. Drugs taken in the previous week had their barcodes scanned and Anatomical Therapeutic Chemical (ATC) codes were obtained.

### The CHRIS kidney questionnaire

To identify different kidney diseases, we developed a dedicated questionnaire based on typical diagnoses and previous experience from a smaller study in the same district.^16,17^ The CHRIS kidney questionnaire was developed in German (the most common language in the district) and Italian, and translated in English. Questionnaires were administered in person by trained study assistants. After the first year of the study, in November 2012, the questionnaire was re-evaluated and improved. The subsequent final version consists of a multiple-choice retrospective questionnaire (**Figure 1**) with a nested structure divided into three sections. Section S1, on specific kidney diseases, started by screening whether a doctor ever told the participant they had a kidney disease of any kind (Q0) and, if yes, asked if they ever had a diagnosis of six specific diseases, namely: glomerulonephritis (Q1), pyelonephritis (Q2), kidney renal artery disease (Q3), hereditary or congenital kidney disease (Q4), kidney stones (Q5), and any other kidney disease not mentioned before (Q6). Each disease had a sub-question on the age at diagnosis and a free-text field. For diseases affecting the glomeruli, we used “glomerulonephritis” as more specific terms would have been difficult to categorize, and because participants frequently reported it in a previous study.^17^ Section S2 was on reduced kidney function (Q7), including questions on dialysis (Q8) and past transplants (Q9). Section S3 dealt with renal surgeries.

**Figure 1.**
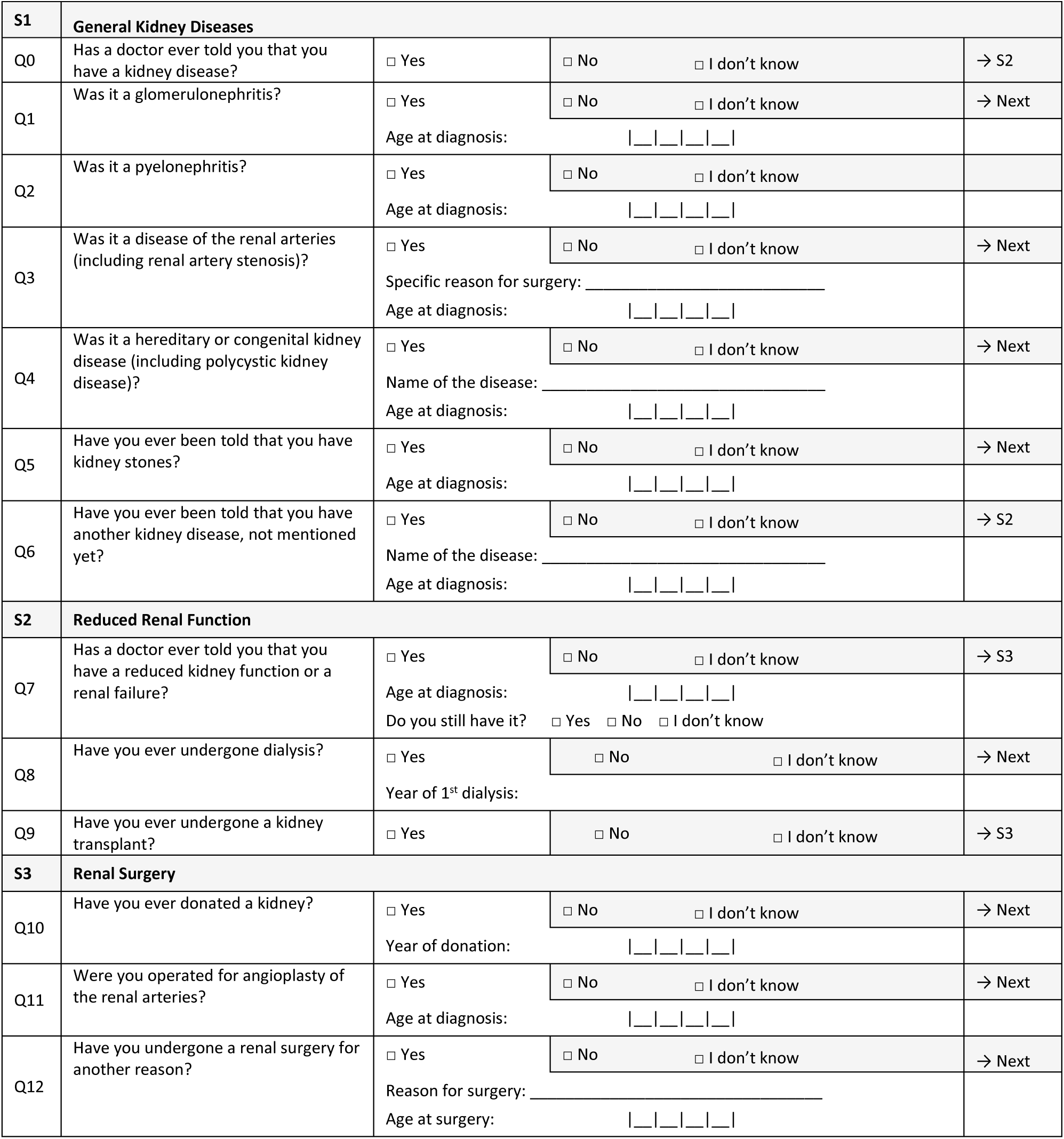
The CHRIS kidney questionnaire.

We analyzed results for all questionnaire items except for dialysis (Q8), transplantation (Q9), and kidney donations (Q10), reported by nobody, and renal artery diseases (Q3, 5 cases) and surgeries (Q11, 2 cases), due to low case numbers.

### Biochemical markers of kidney health

Serum and urine creatinine were measured with the Jaffé method based on Roche Modular PPE and Abbott Diagnostic Architect c16000 instruments.^18^ Urinary albumin was measured with immunonephelometry. Albumin values below the assay limit of detection were set to the limit. We used the race-free 2021 CKD-EPI equation for GFR estimation.^19^ Kidney damage was assessed as the urinary albumin-to-creatinine ratio (UACR). Changes in the measurement assays^18^ were addressed via quantile normalization to the most recent assay.^20^ UACR was log-transformed (logUACR) to mitigate skewness.

### Kidney disease definitions

We defined: increased albuminuria as UACR≥30 mg/g; eGFR-based CKD (CKD_eGFR_) as eGFR<60 mL/min/1.73m^2^; KDIGO-based CKD (CKD_KDIGO_) as UACR>30 mg/g or eGFR<60 mL/min/1.73m^2^, according to KDIGO guidelines;^21^ self-reported CKD (CKD_SR_) as a positive response to Q7 “Has a doctor ever told you that you have a reduced kidney function or a renal failure?”; any CKD (CKD_any_) as either CKD_KDIGO_ or CKD_SR_; and any kidney-related disease as CKD_any_ or any other self-reported kidney disease.

### Comorbidities

Hypertension (HT) was defined as: (i) affirmative answer to “Has a doctor ever said that you have high blood pressure or hypertension?”; (ii) current blood pressure lowering therapy; or (iii) measured systolic or diastolic blood pressure of ≥140 or ≥90 mmHg, respectively. Diabetes mellitus (DM) was defined as: (i) affirmative answer to “Do you have diabetes mellitus?”; (ii) current diabetes medication; or (iii) measured HbA1c ≥6.5%. ATC codes for HT and DM definition are reported in **Supplemental Table S2**.

### Statistical analyses

We excluded 1,698 participants filling the first version of the kidney questionnaire, 6 without kidney questionnaire, and 28 with missing marker measurements, leaving 11,656 participants for statistical analyses (**Supplemental Figure S1**). All prevalence and proportion estimates were standardized to the sex and age structure of the target population via post-stratified relative sampling weights. Prevalence confidence intervals (CIs) were estimated using the Clopper-Pearson method.^22^ For categorical variables with >2 levels, multinomial proportion CIs were estimated with the Wald method.^23^

For each kidney questionnaire item, sex differences were evaluated through age-adjusted logistic regression models with the above-mentioned population sampling weights to account for variations in representativeness. We estimated a pairwise tetrachoric correlation between all questionnaire items. We conducted a first exploratory factor analysis (EFA) to assess the correlation structure using only the questionnaire items. In a second EFA, we introduced increased albuminuria and CKD_eGFR_ to explore the questionnaire’s ability to identify participants with reduced kidney function. To determine the optimal number of factors, we considered the eigenvalues of the correlation matrix that were >1, and looked for sharp breaks in the analysis screeplots. Sensitivity and specificity of each questionnaire item were assessed against CKD_eGFR_, increased albuminuria, and CKD_KDIGO_ as gold standards. We specifically analyzed the association of Q7 responses (‘Yes’, ‘No’, and ‘I do not know’) with eGFR and logUACR levels, by fitting linear models on eGFR and logUACR, adjusting for age and sex and including Q7 responses as a categorical variable. Models were replicated using a six-category exposure variable combining Q7 responses with presence and absence of HT and/or DM.

### Software

All analyses were performed with the R software v4.1.1,^24^ using: the *tetrachoric* and *fa.poly* functions of the ‘psych’ package v2.2.5 for tetrachoric correlation and factor analyses, respectively;^25^ the *confusionMatrix* function of the ‘caret’ package v6.0-93 for sensitivity and specificity analyses;^26^ the *BinomCI* and *MultinomCI* functions of the ‘DescTools’ package v0.99.48 for CI estimation of prevalence and proportions;^27^ the ‘nephro’ package v1.3.0 for GFR estimation.16

## Results

### Study sample characteristics

The 11,656 participants had a median age of 45.5 years (interquartile range, IQR, 30.9-57.3). 53.8% were females, 31.1% had HT, and 3.1% had DM (**Table 1**). eGFR and UACR had median levels of 98.4 mlmL/min/1.73m^2^ (IQR=87.8-108.8) and 5.7 mg/g (IQR=3.8-10.0), respectively, and were slightly negatively correlated (**Figure 4a**). Females had lower eGFR (median 96.2 vs 100.7 ml/min/1.73m^2^) and higher UACR (median 7.2 vs 4.4 mg/g) levels than males.

**Table 1:** Study sample characteristics.

| Characteristics | N (%) or median (IQR) |  |  |
| --- | --- | --- | --- |
|  | Overall | Males | Females |
| Sample size (%) | 11,656 (100) | 5,384 (46.2) | 6,272 (53.8) |
| Age, years, median (IQR) | 45.5 (30.9, 57.3) | 46.1 (31.5, 58.1) | 45.0 (30.4, 56.7) |
| Education, N (%) |  |  |  |
| *Primary school or no title* | 1,225 (10.5) | 475 (8.8) | 750 (12.0) |
| *Lower secondary school* | 1,856 (15.9) | 757 (14.1) | 1,099 (17.5) |
| *Upper secondary school* | 2,708 (23.2) | 1,010 (18.8) | 1,698 (27.1) |
| *Vocational school* | 4,813 (41.3) | 2,759 (51.2) | 2,054 (32.7) |
| *University or higher* | 1,054 (9.0) | 383 (7.1) | 671 (10.7) |
| eGFR, ml/min/1.73m<sup>2</sup>, median (IQR) | 98.4 (87.8, 108.8) | 100.7 (89.9, 111.3) | 96.2 (86.4, 106.6) |
| UACR, mg/g, median (IQR) | 5.7 (3.8, 10.0) | 4.4 (3.2, 7.2) | 7.2 (4.7, 12.1) |
| Diabetes mellitus (DM), N (%) | 359 (3.1) | 168 (3.1) | 191 (3.0) |
| Hypertension (HT), N (%) | 3,623 (31.1) | 1,901 (35.3) | 1,722 (27.5) |
| At least one comorbidity (DM or HT) | 3,712 (31.8) | 1,932 (35.9) | 1,780 (28.4) |

Abbreviations: IQR, interquartile range;

### Analysis of the CHRIS kidney questionnaire

Analysis of the questionnaire showed 743 (6.6%) participants reporting diagnoses of one kidney disease and 183 (1.6%) two or more kidney diseases, with significant sex differences (**Table 2**). Pyelonephritis (3.0%), kidney stones (2.9%), and other unspecified kidney diseases (1.9%) were the most reported. Compared to males, females reported less kidney stones (odds ratio, OR=0.7, 95%CI: 0.5-0.8) and renal surgeries (OR=0.6, 95%CI: 0.4-0.9), and more pyelonephritis (OR=8.7, 95%CI: 6.2-12.3). Years from diagnosis showed disease-dependent distributions (**Table 2**; **Supplemental Figure S2**): the median time from diagnosis was 30 years for glomerulonephritis (IQR=11-43), 29 years for pyelonephritis (IQR=15-39) and 26 years for congenital kidney diseases (IQR=19-32). Kidney stones (median=14, IQR=6-26), other kidney diseases (median=10, IQR=3-24), and reduced kidney function or renal failure (median=10, IQR=3-27) were diagnosed more recently.

**Table 2:**
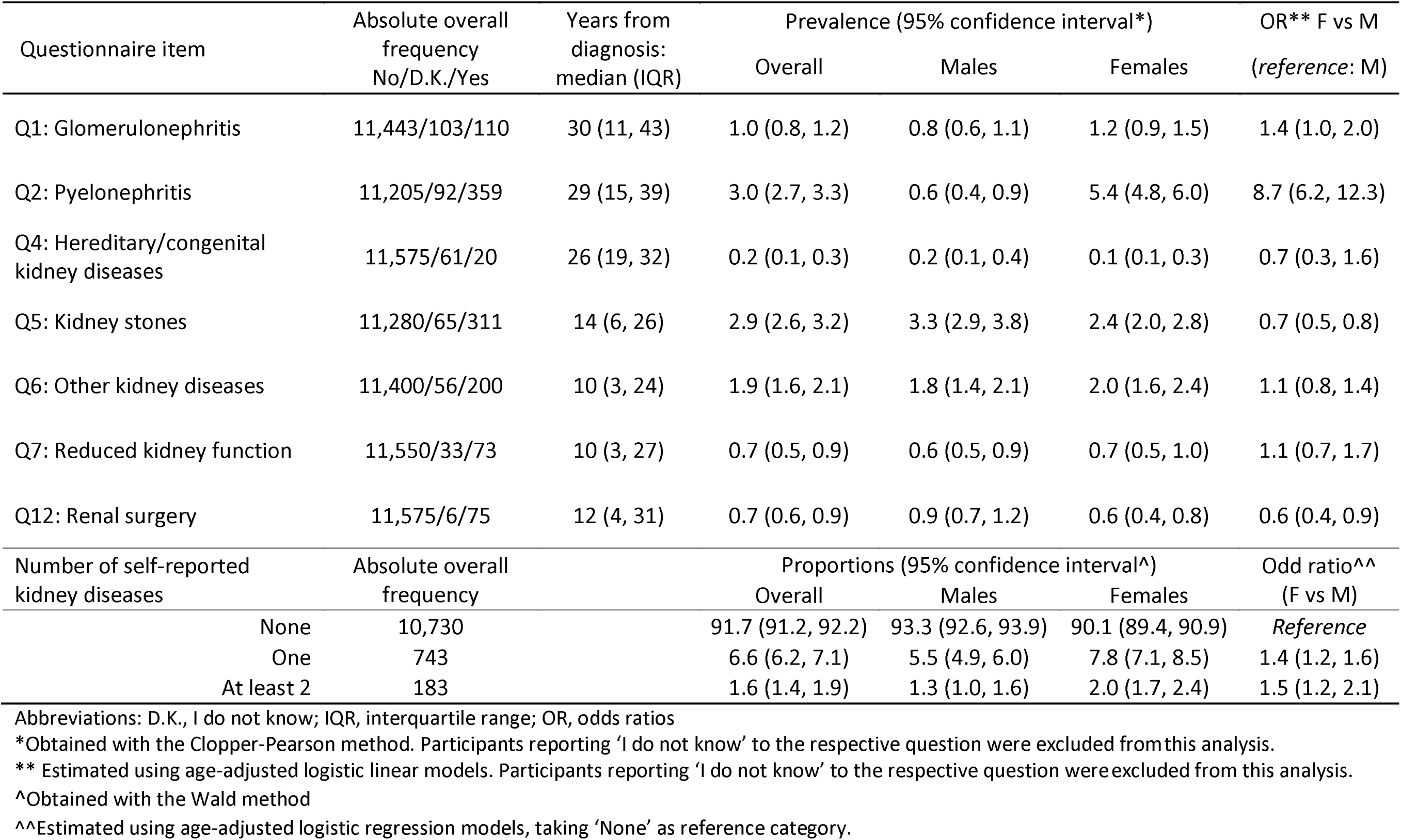
Overall and sex-stratified lifetime prevalence of self-reported kidney diseases calibrated to the general population distribution. Included are also distributions of the time from diagnosis and sex-associated risks of reporting.

All questionnaire items displayed low sensitivity and high specificity in identifying CKD_eGFR_ or increased albuminuria (**Supplemental Table S1**). For instance, reporting a diagnosis of any kidney disease (Q0) showed sensitivity and specificity of 0.27 and 0.92 for CKD_eGFR_, and 0.13 and 0.92 for increased albuminuria, respectively. Q7, querying on CKD diagnosis, exhibited a sensitivity of 0.08 and a specificity of 0.99 for CKD_eGFR_, and 0.02 and 0.99 for increased albuminuria. Factor analysis of the questionnaire items identified one single meaningful factor named ‘general kidney health status’, to which many items contributed substantially equally (**Figure 2a**). When including increased albuminuria and CKD_eGFR_ in the factor analysis, two distinct factors emerged: one was defined as ‘reduced kidney function’, represented by the proximity between increased albuminuria and CKD_eGFR_ with Q7_;_ the second was defined as ‘any other kidney disease’ and corresponded to the combination of all other items except Q7 (**Figure 2b**).

**Figure 2.**
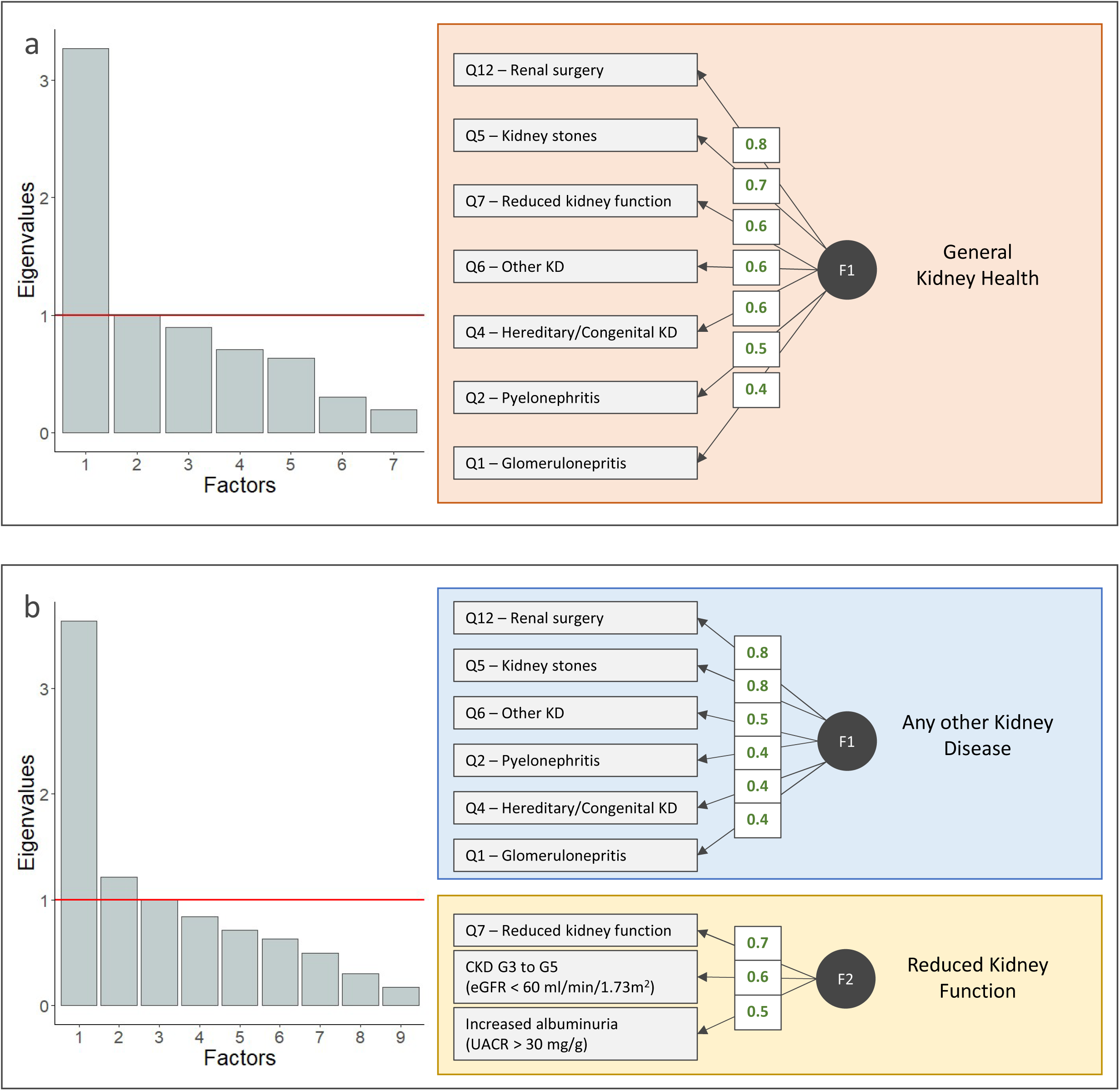
Factor analysis results. **Panel A**: scree plot and graphical summary with component loadings based on the questionnaire only. **Panel B**: scree plot and graphical summary with component loadings based on both the questionnaire and the marker-based diagnoses.

Participants responding ‘I do not know’ to Q7 exhibited eGFR and UACR distributions similar to those responding ‘Yes’ and significantly different from those responding ‘No’ (**Figure 3**): compared to those responding ‘No’, those responding ‘I do not know’ had lower mean eGFR of -10.6 mL/min/1.73m^2^ (95%CI: -14.3, -6.9) and higher mean logUACR of 0.1 log(mg/g) (95%CI: -0.1, 0.4), and those responding ‘Yes’ had lower mean eGFR of -11.7 (95%CI: -14.2, -9.3) ml/min/1.73m^2^ and higher mean logUACR of 0.5 (95%CI: 0.3, 0.7) log(mg/g). Regression models on eGFR and logUACR against combinations of HT, DM, and Q7 responses, showed that, in the presence of HT or DM, those responding ‘I do not know’ had a profile more compatible with the presence rather than absence of CKD; in contrast, in the absence of HT and DM, their values were distributed similar to individuals without CKD (**Supplementary Figure S3a-b**).

**Figure 3.**
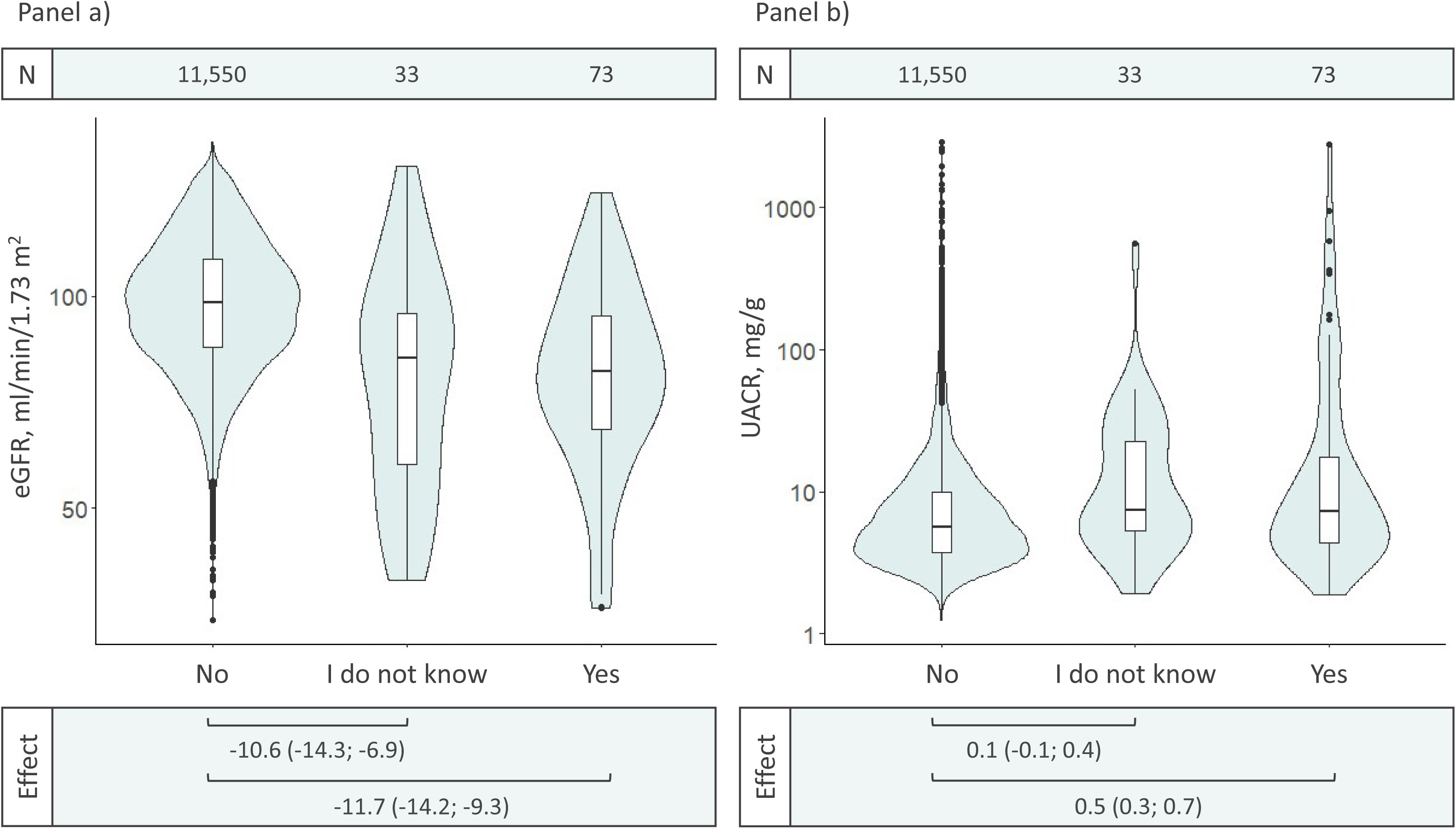
Box-plots and association of responses to the CKD question (Q7) with eGFR and log(UACR). Coefficients of association (effects) and their 95% confidence intervals (CIs) were estimated via age- and sex-adjusted linear regression models.

**Figure 4.**
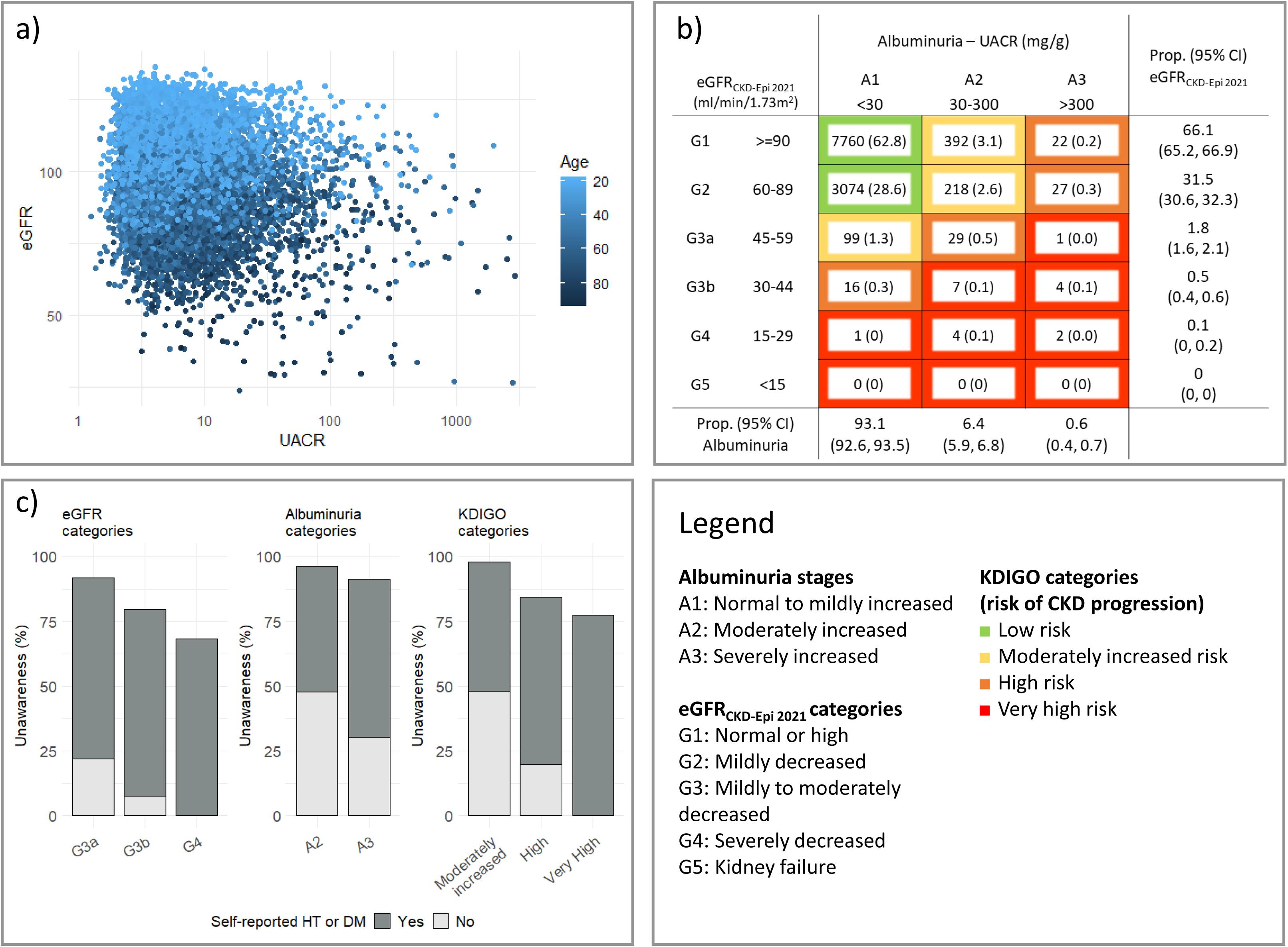
Distribution of CKD and defining traits in the CHRIS study. **Panel A**: Scatter plot of log(UACR) versus eGFR levels, by age of participants. **Panel B**: KDIGO CKD stages calibrated to the general population distribution, with multinomial 95% confidence intervals. **Panel C**: Awareness of CKD across CKD severity stages.

### CKD prevalence and awareness

CKD prevalence varied depending on the definition (**Table 3**). The CKD_KDIGO_ population-representative estimate was 8.59% (95%CI: 8.09%-9.12%) and was driven by increased albuminuria (prevalence=6.95%, 95%CI: 6.49-7.42%) rather than low eGFR levels (CKD_eGFR_ prevalence=2.42%; 95%CI: 2.15%-2.72%). The KDIGO classification (**Figure 4b**) identified no participants in the eGFR stage G5 and a 0.6% prevalence (95%CI: 0.4%-0.7%) of albuminuria stage A3. Overall, 0.3% (95%CI: 0.2%-0.4%) of individuals had a very high risk of CKD progression, incident CVD events and mortality.

**Table 3:**
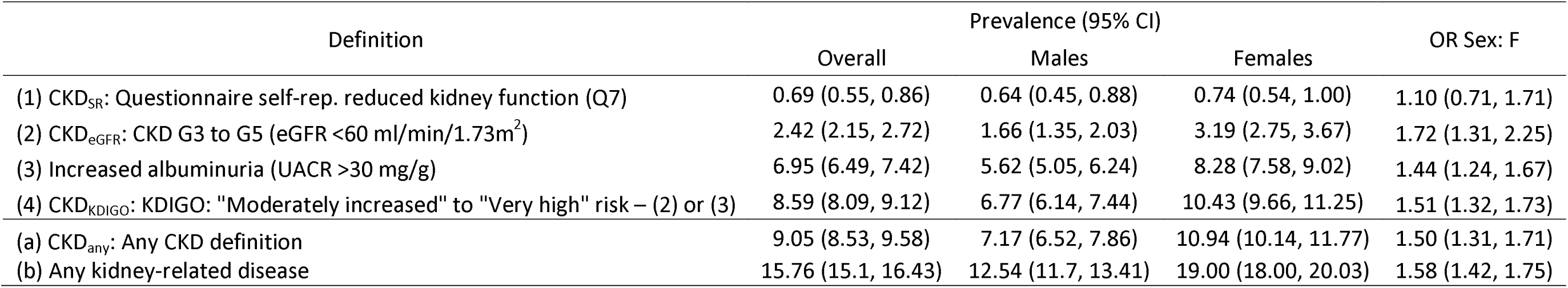
Prevalence of CKD in the Val Venosta/Vinschgau district using different definitions.

| Definition | Prevalence (95% CI) |  |  |  |
| --- | --- | --- | --- | --- |
|  | Overall | Males | Females | OR Sex: F |
| (1) CKD<sub>SR</sub>: Questionnaire self-rep. reduced kidney function (Q7) | 0.69 (0.55, 0.86) | 0.64 (0.45, 0.88) | 0.74 (0.54, 1.00) | 1.10 (0.71, 1.71) |
| (2) CKD<sub>eGFR</sub>: CKD G3 to G5 (eGFR <60 ml/min/1.73m<sup>2</sup>) | 2.42 (2.15, 2.72) | 1.66 (1.35, 2.03) | 3.19 (2.75, 3.67) | 1.72 (1.31, 2.25) |
| (3) Increased albuminuria (UACR >30 mg/g) | 6.95 (6.49, 7.42) | 5.62 (5.05, 6.24) | 8.28 (7.58, 9.02) | 1.44 (1.24, 1.67) |
| (4) CKD<sub>KDIGO</sub>: KDIGO: "Moderately increased" to "Very high" risk – (2) or (3) | 8.59 (8.09, 9.12) | 6.77 (6.14, 7.44) | 10.43 (9.66, 11.25) | 1.51 (1.32, 1.73) |
| (a) CKD<sub>any</sub>: Any CKD definition | 9.05 (8.53, 9.58) | 7.17 (6.52, 7.86) | 10.94 (10.14, 11.77) | 1.50 (1.31, 1.71) |
| (b) Any kidney-related disease | 15.76 (15.1, 16.43) | 12.54 (11.7, 13.41) | 19.00 (18.00, 20.03) | 1.58 (1.42, 1.75) |

In contrast, CKD_SR_ prevalence was as low as 0.69% (95%CI: 0.55%-0.86%). Prevalence of CKD_any_ (CKD_KDIGO_ or CKD_SR_) was 9.05% (95%CI: 8.53%-9.58%). Besides CKD_SR_, any other type of CKD was more prevalent in females than males. We estimated that 15.76% (95%CI: 15.10%-16.43%) of the district’s adult population had experienced at least one kidney disease of any type.

Of the 822 individuals with CKD_KDIGO_, only 31 reported CKD_SR_. Thus, the standardized proportion of individuals with CKD_KDIGO_ who are ‘unaware’ of having CKD is 95.3% (95%CI: 93.8%-96.5%). The unawareness decreased with increasing disease severity (**Figure 4c**; **Supplemental Table S3**), although heterogeneously across outcomes: unawareness was >90% among albuminuria stage A3 individuals and 79.6% among those with eGFR stage G4 (**Supplemental Table S3**). Most unaware individuals reported having HT or DM. This occurred in approximately two-thirds of the individuals with albuminuria stage A3 and 100% of those with eGFR stage G4 (**Figure 4c**).

## Discussion

Our investigation of CKD and kidney health in the Val Venosta/Vinschgau district (South Tyrol, Italy), combining self-reported data and biomarkers, reveals a CKD prevalence of approximately 9%. It offers firsthand experience with implementing a dedicated kidney health questionnaire outside clinical settings and quantifies lack of CKD awareness in the general population.

CKD prevalence in the district was lower than the 12.7% to 15.5% reported by some Italian population cohort studies (INCIPE^6^ and SardiNIA^28^, respectively), yet higher than others,^7,8^ while generally matching figures from other Western-European countries.^5^ Sharing geographic and cultural proximity with the CHRIS study, the KORA study reported prevalence of CKD_eGFR_ and CKD_KDIGO_ of 9.7% and 16.0%, respectively.^29^

To our knowledge, no prior study has systematically investigated the lifetime prevalence of a broad spectrum of self-reported kidney diseases in the general population. We implemented the novel CHRIS kidney questionnaire to address this gap. Study participants reporting at least one kidney disease exceeded 7%. The most reported condition was pyelonephritis (3%), with an almost ninefold higher likelihood of reporting for females than males, consistent with literature indicating a higher risk of urinary tract infections, including pyelonephritis, in females.^30,31^ In addition to clinical reasons, the higher self-reported prevalence of both pyelonephritis and glomerulonephritis in females surveys could also reflect some level of under-reporting among males.^32^ Among the other conditions, congenital diseases were reported by 0.2% of the sample, and kidney stones by just less than 3%.

As expected, all conditions presented a high-to-nearly-perfect specificity and low-to-null sensitivity regarding CKD_KDIGO_, reflecting that the listed diseases do not necessarily affect eGFR and UACR, except in the long term. Questions on dialysis, transplantation, and renal surgeries yielded no insight, with few or no participants reporting such conditions. This suggests caution when selecting survey questions for the general population, as their inclusion may cause substantial effort for both the interviewer and the interviewee, with minimal or no benefit, potentially compromising response accuracy.

The questionnaire effectively discriminated between kidney pathologies responding to a specific diagnosis and identifiable symptoms, recognizable by both patients and physicians, and CKD, which may progress latently by reduction of kidney function levels, albeit undiagnosed. This finding is reflected by factor analysis results including both CKD_SR_ and measured kidney function markers: CKD_SR_ clustered well with measured eGFR and UACR levels, while all other questionnaire items clustered apart. This observation supports the importance of implementing kidney health questionnaires for population studies as they capture disease domains not typically identified by routinely measured markers.

A substantial majority of individuals with CKD_KDIGO_ were unaware of their condition. This aligns with recent literature reporting rates of underdiagnosis between 61.6% in the USA and 95.5% in France.^11^ We acknowledge a potential slight CKD_KDIGO_ prevalence overestimation due to inclusion of acute events coming from the cross-sectional nature of the CHRIS study, with biochemical markers measured at a single instance and the impossibility to obtain a second measure at the recommended three-month timespan.^33^ However, that would not change substantially the figure of unawareness. Our results could reflect a lack of renal health education among participants, stemming from limited attention to CKD by health providers. It is noteworthy that CKD was included in the new essential assistance levels, a set of essential healthcare services and standards established by the Italian government, since November 2017.^34^ Therefore, many healthcare authorities might not yet have implemented measures to raise attention towards CKD during the 2011-2018 data collection period. As the results show, awareness was poor even in the presence of diagnosed diabetes or hypertension. This could imply suboptimal compliance with guidelines recommending periodic creatinine assessments for hypertensive^35^ or diabetic^36^ individuals. Alternatively, individuals diagnosed with diabetes or hypertension might not receive a formal CKD diagnosis due to reduced kidney function being assumed as a standard comorbidity. Another relevant aspect of CKD awareness or diagnosis in our analyses is that CKD_KDIGO_ was more common in females, while CKD_SR_ prevalence was similar between sexes. This indicates higher under-reporting or unawareness in females than males, consistent with literature indicating a lower likelihood of CKD diagnosis, monitoring, and management in women.^37,38^ Consequently, while a certain degree of underdiagnosis is generally expected for a silent disease like CKD, even in electronic health records,^9^ addressing the issue of CKD underdiagnosis becomes imperative also as a way to reduce gender inequalities in healthcare provision.

Strengths of our analyses are the large sample size, the possibility to calibrate results to the target population to obtain representative estimates and mitigate selection bias, and the simultaneous availability of serum creatinine and UACR, which allowed to better gauge the KDIGO criteria.

There are also several limitations. Beyond the already discussed cross-sectional nature of the design, precluding the possibility of repeated assessments, measured GFR was unavailable, nor was cystatin C that would have provided more reliable GFR estimates. However, assessing eGFR through the creatinine-based CKD-EPI equation allowed comparison to most available population-based studies. Moreover, due to logistic and motivational issues, population-based studies like CHRIS suffer the difficulty of recruiting individuals at older ages or with severe chronic conditions. Indeed, the most severe CKD stages are not represented in the CHRIS study, suggesting underestimation of our figures. In addition, despite the revisions made to the questionnaire, we cannot override poor question wording, potentially leading to underestimation and perhaps even misclassification. Furthermore, we cannot exclude participants’ confusion or forgetfulness, not least because some diagnoses dated significantly back in time, leading to potential recall bias. Moreover, questions required the use of technical terminology, unfamiliar to the participants, also because clinicians typically communicate with patients by explaining symptoms and consequences rather than using medical jargon, difficult for the average patient to comprehend. On the other hand, terms like glomerulonephritis, for instance, are just generic, as they include various specific subpathologies, like IgA nephropathy, lupus nephritis or membranous nephropathy.^13^ However, including these specific terms might not be worthwhile as these are rarer conditions and may represent an unfamiliar vocabulary for study participants. Therefore, it seems reasonable to assume that the estimated prevalence of pyelonephritis and glomerulonephritis is an underestimation of the real presence in the reference population. Linkage to electronic health records might overcome these limitations, however this was not yet implemented in the CHRIS study. While imperfect, the CHRIS kidney questionnaire seems to us a helpful step towards identifying disease presence and raising awareness.

In conclusion, understanding the coverage of CKD diagnosis and the extent of individuals’ awareness about their CKD status and the associated risk factors is crucial in promoting early diagnosis and effective management. Enhancing awareness can empower individuals to adopt healthier lifestyles and engage in proactive healthcare behaviors, thus reducing CKD burden on individuals and healthcare systems. While producing population-representative estimates of CKD prevalence in the study area, the CHRIS study provides a rare example of a kidney health questionnaire for general population studies. The extremely high degree of CKD awareness reflects the international context, reinforcing the need for more accurate tools to measure kidney diseases in the general population.

## Data Availability

The data used in the current study can be requested with an application to at the Eurac Research Institute for Biomedicine.

## Acknowledgments

The CHRIS study is conducted in collaboration between the Eurac Research Institute for Biomedicine and the Healthcare System of the Autonomous Province of Bolzano-South Tyrol. Investigators thank all study participants, the general practitioners of the Val Venosta/Vinschgau district, the staff of the Silandro/Schlanders hospital and of the Autonomous Province of Bolzano-South Tyrol Healthcare System for their support and collaboration, the CHRIS Biobank personnel, and all Institute for Biomedicine colleagues who contributed to the study. Investigators also thank the study assistants and nurses (Karin Bystrianska, Brunhilde Grasser, Roselinde Gunsch, Benedikta Linter, Liane Parth, Susanne Saewert, Renate Telser) for their helpful feedback on the conduction of the kidney questionnaire interviews that helped to understand the difficulties related to the use of specific questions and in the interpretation of non-responses. Extensive acknowledgement is reported at: https://www.eurac.edu/en/institutes-centers/institute-for-biomedicine/pages/acknowledgements. CHRIS Bioresource Research Impact Factor (BRIF) code: BRIF6107.

## Author contribution

Conceptualization: GB, LC, GG, CP, MEZ

Data curation: GB, LB, MG

Formal Analysis: GB

Investigation: GB, LC, RM, MEZ, CP

Methodology: GB, LC, RM, MG, CP, MEZ

Project administration: PPP, RL, CP

Resources: PPP, RL, MG, CP

Funding acquisition: PPP, EH, CP

Supervision: LC, MEZ, CP

Visualization: GB

Writing – original draft: GB, CP

Writing – review & editing: All

## Ethics approval and consent to participate

The Ethics Committee of the Healthcare System of the Autonomous Province of Bolzano-South Tyrol approved the CHRIS baseline protocol on 19 April 2011 (21-2011). The study conforms to the Declaration of Helsinki, and with national and institutional legal and ethical requirements. All participants included in the analysis gave written informed consent.

## Data sharing statement

CHRIS study data and samples can be requested for research purposes by submitting a dedicated request to the Eurac Research Institute for Biomedicine Access Committee at.

