## Supplemental for "Development and evaluation of a kidney health questionnaire and estimates of chronic kidney disease prevalence in the Cooperative Health Research In South Tyrol (CHRIS) study"

**Supplementary Material**

### **Table S1**: Specificity and sensitivity for each questionnaire item and kidney dichotomous outcomes

| Questionnaire items | Absolute overall frequency No/Yes | Sensitivity/Specificity | | |
| --- | --- | --- | --- | --- |
|  |  | CKD_eGFR_ | Increased albuminuria | CKD_KDIGO_ |
| Q0: Any kidney disease | 10,687/921 | 0.27/0.92 | 0.13/0.92 | 0.15/0.93 |
| Q1: Glomerulonephritis | 11,443/110 | 0.05/0.99 | 0.02/0.99 | 0.02/0.99 |
| Q2: Pyelonephritis | 11,205/359 | 0.09/0.97 | 0.04/0.97 | 0.05/0.97 |
| Q4: Hereditary/congenital kidney diseases | 11,575/20 | 0.01/1.00 | 0.00/1.00 | 0.00/1.00 |
| Q5: Kidney stones | 11,280/311 | 0.09/0.97 | 0.04/0.97 | 0.05/0.98 |
| Q6: Other kidney diseases | 11,400/200 | 0.09/0.98 | 0.04/0.98 | 0.05/0.98 |
| Q7: Reduced kidney function | 11,550/73 | 0.08/0.99 | 0.02/0.99 | 0.03/1.00 |
| Q12: Renal surgery | 11,575/75 | 0.04/0.99 | 0.01/0.99 | 0.01/0.99 |

### **Table S2**: Anatomical therapeutic chemical (ATC) codes used to identify self-reported medications for hypertension and diabetes

| Type of medication | Therapy ( ATC code) |
| --- | --- |
| Blood pressure lowering drugs | Antihypertensives (C02), Diuretics (C03), Peripheral vasodilators (C04), Beta blocking agents (C07), Calcium channel blockers ( C08), Agents acting on the renin-angiotensin system (C09) |
| Drugs used in diabetes | Insulins and analogues (A10A), Blood glucose lowering drugs (A10B) |

### **Table S3**: Calibrated proportion of participants within the different stages of CKD and multinomial 95% Confidence Intervals

| Subclinical CKD stages | Overall (N=11,656) | | | | In those not reporting HT and DM (N=8,832) | In those reporting HT and DM (N=2,824) |
| --- | --- | --- | --- | --- | --- | --- |
|  | N | | Prop. (95% CI) | | Prop. (95% CI) | Prop. (95% CI) |
| CKD eGFR Stages |  | |  | |  |  |
| *G1* | 8174 | | 66.1 (65.2, 66.9) | | 75.1 (74.2, 76.0) | 42.6 (40.9, 44.3) |
| *G2* | 3319 | | 31.5 (30.6, 32.3) | | 24.3 (23.4, 25.2) | 50.3 (48.5, 52.0) |
| *G3a* | 129 | | 1.8 (1.6, 2.1) | | 0.6 (0.4, 0.7) | 5.1 (4.3, 5.8) |
| *G3b* | 27 | | 0.5 (0.4, 0.6) | | 0.1 (0.0, 0.1) | 1.7 (1.2, 2.1) |
| *G4* | 7 | | 0.1 (0.0, 0.2) | | -- | 0.4 (0.2, 0.6) |
| *G5* | 0 | | -- | | -- | -- |
| Albuminuria Stages |  | |  | |  |  |
| *A1* | 10950 | | 93.1 (92.6, 93.5) | | 95.5 (95.1, 96) | 86.6 (85.5, 87.8) |
| *A2* | 650 | | 6.4 (5.9, 6.8) | | 4.2 (3.8, 4.7) | 12.0 (10.8, 13.1) |
| *A3* | 56 | | 0.6 (0.4, 0.7) | | 0.2 (0.1, 0.4) | 1.4 (1.0, 1.8) |
| KDIGO categories |  | |  | |  |  |
| *Low risk* | 10834 | | 91.4 (90.9, 91.9) | | 94.9 (94.5, 95.4) | 82.2 (80.9, 83.5) |
| *Moderate increased risk* | 709 | | 7.0 (6.6, 7.5) | | 4.7 (4.3, 5.2) | 13.1 (11.9, 14.2) |
| *High risk* | 94 | | 1.2 (1.0, 1.4) | | 0.3 (0.2, 0.5) | 3.5 (2.9, 4.1) |
| *Very high risk* | 19 | | 0.3 (0.2, 0.4) | | -- | 1.2 (0.8, 1.6) |
| CKD unawareness | Overall (N=11,656) | | | | In not reporting HT and DM (N=8,832) | In reporting HT and DM (N=2.824) |
|  | N |  | | Unawareness (95% CI) | Unawareness (95% CI) | Unawareness (95% CI) |
| CKD eGFR Stages |  | |  | |  |  |
| *G1* | 8136 | | 99.5 (99.3, 99.7) | | 99.5 (99.3, 99.7) | 99.5 (99.0, 99.8) |
| *G2* | 3271 | | 98.5 (98.0, 98.8) | | 98.8 (98.3, 99.3) | 98.0 (97.2, 98.6) |
| *G3a* | 118 | | 91.8 (87.3, 95.1) | | 96.0 (86.1, 99.5) | 90.6 (85.0, 94.6) |
| *G3b* | 21 | | 79.6 (67.0, 89.0) | | 100.0 (43.5, 100.0) | 77.9 (64.5, 88.0) |
| *G4* | 4 | | 68.3 (35.9, 91.2) | | -- | 68.3 (35.9, 91.2) |
| *G5* | 0 | | -- | | -- | -- |
| Albuminuria Stages |  | |  | |  |  |
| *A1* | 10866 | | 99.1 (99.0, 99.3) | | 99.4 (99.2, 99.5) | 98.5 (98.0, 98.9) |
| *A2* | 634 | | 96.2 (94.6, 97.5) | | 99.0 (97.3, 99.8) | 93.7 (90.8, 95.9) |
| *A3* | 50 | | 91.3 (81.8, 96.8) | | 95.7 (76.9, 99.9) | 89.3 (76.5, 96.5) |
| KDIGO categories |  | |  | |  |  |
| *Low risk* | 10759 | | 99.3 (99.1, 99.4) | | 99.4 (99.2, 99.5) | 98.9 (98.4, 99.3) |
| *Moderate increased risk* | 697 | | 98.0 (96.8, 98.9) | | 98.6 (96.9, 99.5) | 97.4 (95.5, 98.7) |
| *High risk* | 80 | | 84.5 (77.4, 90.0) | | 96.9 (82.8, 99.9) | 81.3 (72.8, 88.0) |
| *Very high risk* | 14 | | 77.4 (61.1, 89.3) | | -- | 77.4 (61.1, 89.3) |

### **Figure S1**: Flowchart of the study sample

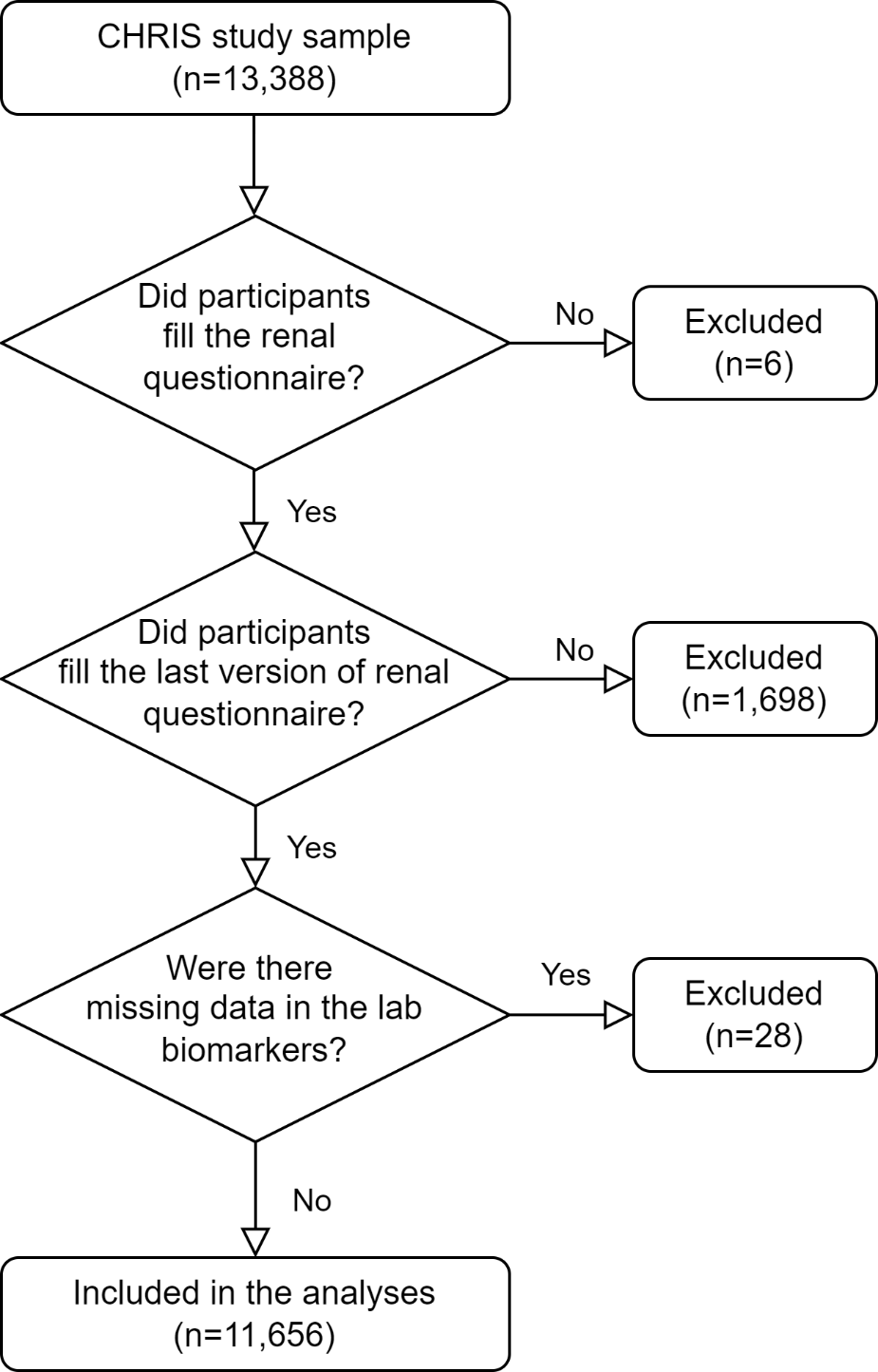

### **Figure S2**: Distribution of time from diagnosis (years) for various self-reported kidney diseases

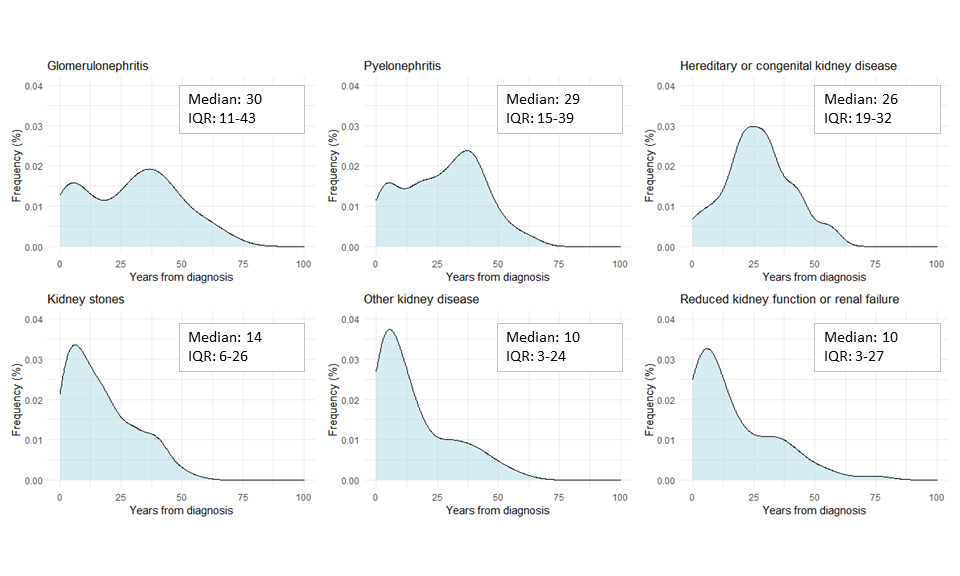

### **Figure S3a:** Boxplots, β coefficients and 95% Confidence Intervals (95%CI) of the associations between self-reported renal insufficiency or hypertension/diabetes and levels of eGFR. All models are adjusted for age and sex.

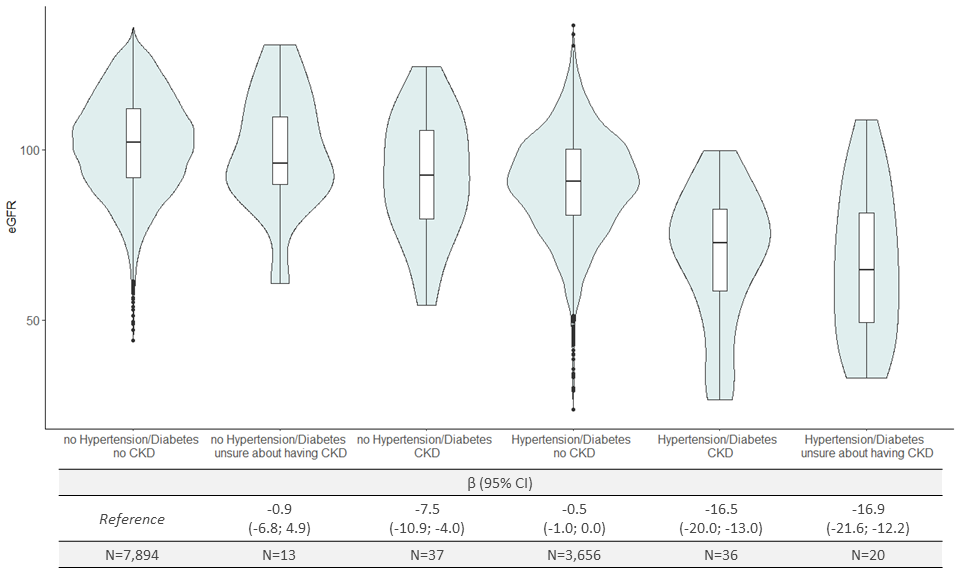

### **Figure S3b:** Boxplots, β coefficients and 95% Confidence Intervals (95%CI) of the associations between self-reported renal insufficiency or hypertension/diabetes and levels of UACR. All models are adjusted for age and sex.

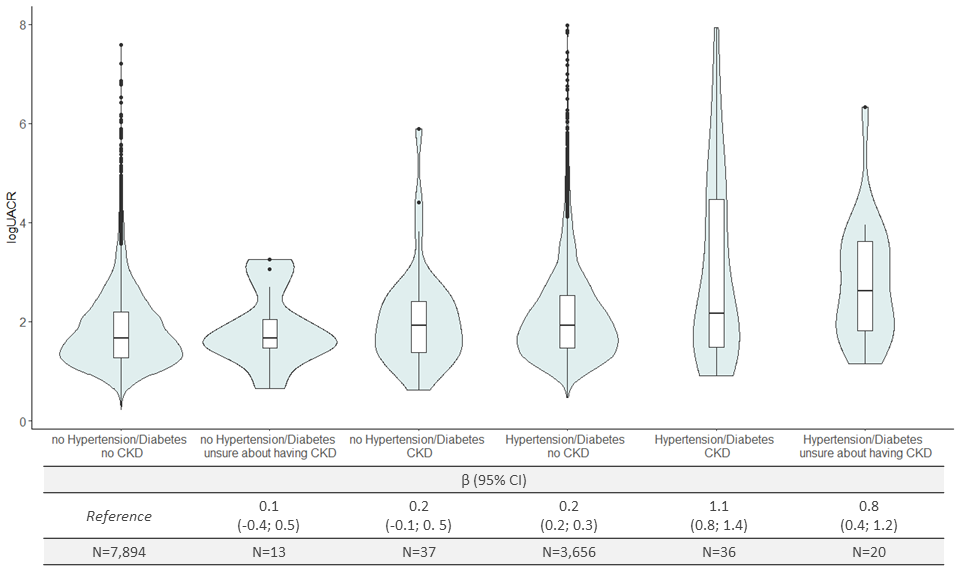
